# Probabilistic comparison of gray and white matter coverage between depth and surface intracranial electrodes in epilepsy: a patient-specific modeling and empirical study

**DOI:** 10.1101/2021.08.04.21261603

**Authors:** Daria Nesterovich Anderson, Chantel M. Charlebois, Elliot H. Smith, Amir M. Arain, Tyler S. Davis, John D. Rolston

## Abstract

**Objective:** The objective of this study is to quantify the coverage of gray and white matter during intracranial electroencephalography in a cohort of epilepsy patients with surface and depth electrodes.

**Methods:** We included 65 patients with strip electrodes (n=12), strip and grid electrodes (n=24), strip, grid, and depth electrodes (n=7), or depth electrodes only (n=22) from the University of Utah spanning 2010-2020. Patient-specific imaging was used to generate probabilistic gray and white matter maps and atlas segmentations. The gray and white matter coverage was quantified based on spherical volumes centered on electrode centroids, with radii ranging from 1-15 mm, along with detailed finite element models of local electric fields

**Results:** Gray matter coverage was highly dependent on the chosen radius of influence (RoI). Using a 2.5 mm RoI, depth electrodes covered more gray matter than surface electrodes; however, surface electrodes covered more gray matter at RoI larger than 4 mm. White matter coverage was greatest for depth electrodes at all RoIs, which is noteworthy for studies involving stimulation mapping. Depth electrodes were able to record significantly more gray matter from the amygdala and hippocampus than subdural electrodes.

**Significance:** This study provides the first probabilistic analysis to quantify gray and white matter coverage for multiple categories of intracranial recording configurations. Depth electrodes may offer increased per contact coverage of gray matter over other recording strategies if the desired signals are local to the contact, while subdural grids and strips can sample more gray matter if the desired signals are more diffuse.

## Introduction

Intracranial monitoring for epilepsy is approaching a century of clinical use [1,2], and to this day, a debate over the benefits and drawbacks for surface electrodes (i.e., subdural strips and grids) or depth electrodes (i.e., stereo-electroencephalography, SEEG) continues. The use of a specific intracranial recording modality is often dependent on physician preference, with some broad trends apparent from country to country [3]. In the United States specifically, the use of SEEG has been increasing in prevalence [4]. A possible concern in adopting exclusive use of depth electrodes is the potential difference in gray matter coverage when compared to the apparent cortical coverage using classic subdural electrodes.

A common assertion is that subdural electrodes cover more gyral gray matter, while depth electrodes cover more sulcal gray matter [5]. Subdural grids may also be preferred due to their fixed spatial relationships across electrodes and stereotyped coverage of the cortex. While subdural grids cover what appear to be neighboring brain regions, as has been previously reported [3,5], the amount of gray matter buried in sulci complicates the notion of contiguous coverage. That is, even though grids have fixed 2-dimensional arrangements, the brain beneath them does not. Though adequate coverage of gray matter is critical in choosing a recording modality, the predicted location of the epileptic focus, known as the localization hypothesis, heavily plays into the decision into which modality to use. If the seizure onset zone was thought to arise from a deep lesion or if there were a reason to record bilaterally, the use of depth electrodes might be favored [3,6]. Risk profiles differ between both approaches, and this also plays a role in choosing which modality to use [7–12]. Finally, each method of intracranial monitoring may have advantages over others, yet the decision to use a certain modality over another at many US institutions may have more to do with preferred clinical practice at a given center [3].

While many factors may guide the choice to use a certain intracranial monitoring technique, if one modality demonstrates superior gray matter coverage, it may increase the likelihood that the epileptic zone would be appropriately sampled. To elucidate these differences, we aimed to quantify the coverage of gray and white matter and region-specific coverage for subdural and depth electrode cases through a probabilistic and patient-specific computational analysis.

## Methods

A total of 65 patients were included in our study, falling into the following categories: 1) patients using only subdural strip electrodes (S), 2) patients using a combination of subdural strip electrodes and subdural grid electrodes (S + G), 3) patients using a combination of subdural strip, subdural grid, and depth electrodes (S + G + D), 4) and patients using only depth electrodes (D) (i.e., SEEG). Throughout this study, we will use the term ‘electrode’ to reference the individual contacts on depth leads or strip and grids arrays. Additionally, any reference to surface electrodes are referring to intracranial surface (epicortical) electrodes, not scalp electrodes. All cases were performed at the University of Utah hospital between 2010-2020 under two neurosurgeons. The study was approved by the University of Utah Institutional Review Board.

For patients who had undergone intracranial monitoring multiple times at the University of Utah for further diagnosis of the seizure onset zone, only the first invasive electrode placement was considered in this analysis. Table 1 summarizes the relevant patient characteristics, hemispheric coverage, and the average number of strip, grid, and depth electrodes in each category. Figure 1A provides an overview of the patient processing pipeline to generate gray and white matter coverage volumes for all types of intracranial modalities.

**Table 1.** Overview of patient characteristics and the use of strip, grid, and depth electrodes. Ages and electrode counts are reported as mean (standard deviation).

|  | All Patients | Strip | Strip + Grid | Strip + Grid + Depth | Depth |
| --- | --- | --- | --- | --- | --- |
| Total | 65 | 12 | 24 | 7 | 22 |
| Female | 33 | 6 | 15 | 4 | 8 |
| Male | 32 | 6 | 9 | 3 | 14 |
| Age | 33.6 (10.2) | 30.6 (6.9) | 34.8 (11.0) | 32.4 (11.0) | 34.2 (10.8) |
| Epilepsy Onset Age | 14.4 (11.2) | 14.4 (9.9) | 12.3 (11.2) | 18.0 (12.0) | 15.7 (11.9) |
| Bilateral | 38 | 11 | 5 | 2 | 20 |
| Unilateral | 27 | 1 | 19 | 5 | 2 |
| Total Electrodes | 87.6 (20.1) | 74.6 (16.2) | 86.4 (22.8) | 98.3 (13.5) | 92.8 (18.5) |
| Strip Electrodes | 25.0 (28.5) | 74.6 (16.2) | 23.7 (15.3) | 22.6 (8.3) | -- |
| Grid Electrodes | 30.1 (34.7) | -- | 62.7 (21.5) | 64.3 (17.6) | -- |
| Depth Electrodes | 32.4 (44.4) | -- | -- | 11.4 (5.9) | 92.8 (18.5) |

**Figure 1.**
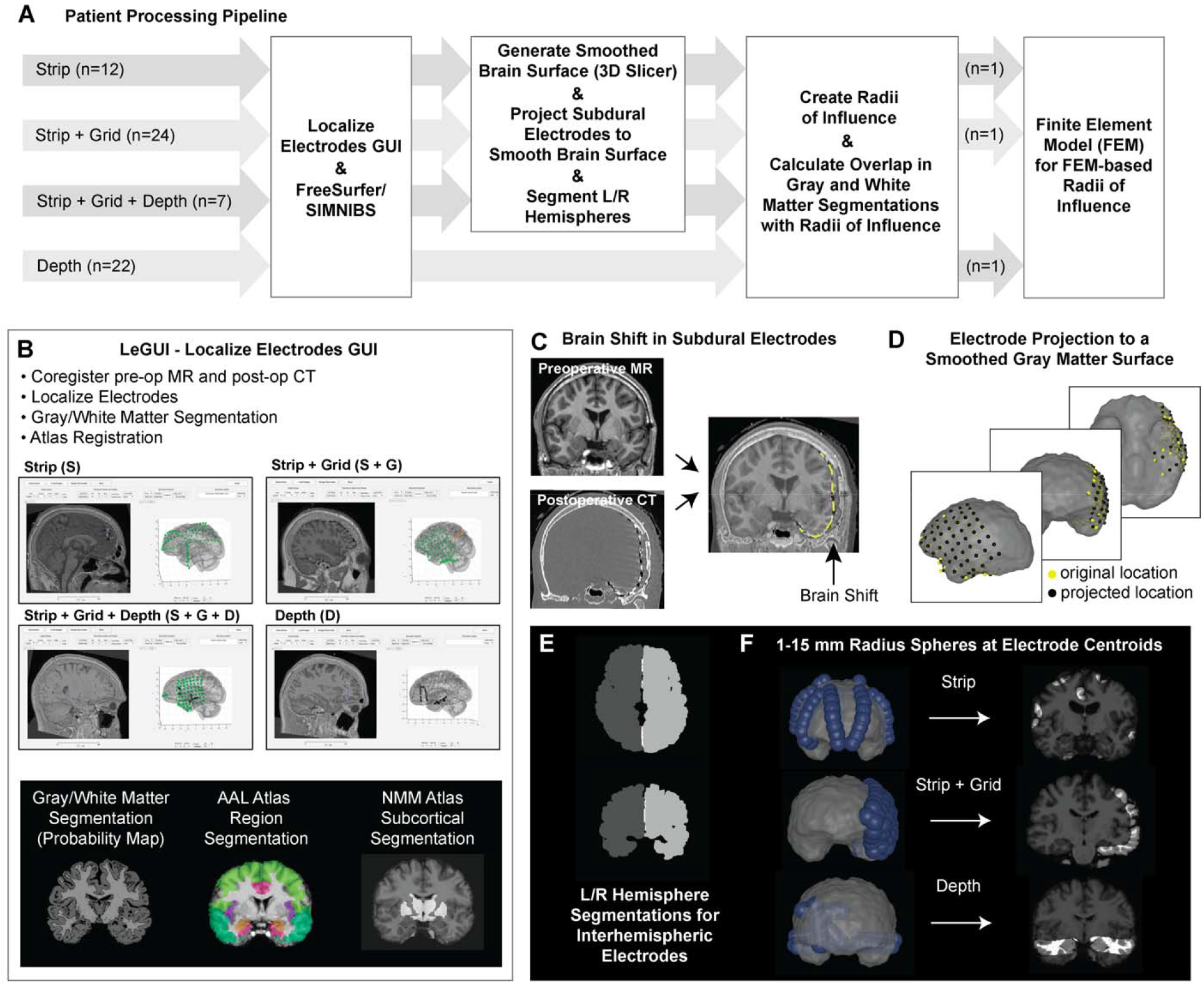
Overview of Methods. **A**. The processing pipeline used patient specific imaging. All patients were processed through LeGUI, FreeSurfer, and SIMNIBS, while only surface electrode patients received extra processing to project the electrodes to the smoothed gray matter surface as well as left and right hemisphere segmentation for interhemispheric contacts. Overlap of gray and white matter segmentations with RoIs were used to calculate volumes of gray and white matter coverage. A subset of patients were used to run FEM-based RoIs. Reported sample sizes indicate the number of patients included in each implant category. **B**. Electrode localization was performed in the freely available LeGUI software to mark strip, grid, and depth electrodes across a cohort of 65 patients. LeGUI automatically processes probabilistic gray and white matter segmentations and numerous atlas registrations including the AAL and NMM atlases. **C**. Substantial brain shift is frequently observed in subdural electrodes cases during surgical implantation. After image co-registration, the subdural electrodes localized from the CT may appear inside of the brain (yellow) rather than on top of the cortical surface. **D**. All subdural electrodes were projected (yellow – original location, black – projected location) onto the smooth gray matter surface according to Hermes et al., 2010. **E**. In patients with interhemispheric subdural electrodes, the left and right hemisphere segmentations were created in FreeSurfer so that volumes would be restricted to the hemisphere from which the electrode recorded neural activity. **F**. Spheres at fixed radii from 1-15 mm at 0.5 mm steps were placed at the contact centroids. 10 mm RoIs are shown in blue. Overlap of spherical volumes with gray and white matter segmentations quantify the amount of gray and white matter coverage for different modalities.

Preoperative structural MRI and postoperative CT scans were linearly coregistered using LeGUI (<u>L</u>ocalize <u>E</u>lectrodes <u>GUI</u>, https://github.com/Rolston-Lab/LeGUI) featuring MATLAB (MathWorks, Natick, Massachusetts) and the SPM12 software package (Statistical Parametric Mapping, https://www.fil.ion.ucl.ac.uk/spm/) (Figure 1B). Gray and white matter segmentations, generated as probability maps of gray and white matter regions, were computed for each structural MR image in LeGUI. Patient structural imaging was nonlinearly registered to the Neuromorphometrics (NMM) brain atlas and Macroscopic Parcellations of the AAL brain atlas (Neuromorphometrics, Inc., http://neuromorphometrics.com/, [13,14]). Subcortical regions were isolated from the NMM atlas and include the nucleus accumbens, caudate, pallidum, putamen, thalamus, and ventral diencephalon. Macro-scale brain regions used to compare region-specific gray matter coverage across modalities were taken from the AAL atlas and include the frontal lobe, temporal lobe (neocortex only), amygdala, hippocampus, insular cortex, and cingulate cortex.

Electrode centroids were localized in LeGUI using the CT artifact. For epicortical electrodes, especially subdural grids, there may be significant brain shift in the cortical surface to accommodate placement of the subdural electrodes and CSF leakage; in some cases, the brain surface may shift greater than 1 cm from the original preoperative MR [15]. A simple linear alignment of preoperative MR and postoperative CT images would, therefore, lead to overcalculation of gray matter coverage statistics, given that most electrodes would be incorrectly localized inside the gray matter (Figure 1C). To combat this, we used a well-established projection algorithm [16,17] to bring the centroids of all subdural electrodes to the surface of the smooth gray matter surface (Figure 1D). The smooth gray matter surface was generated from the gray matter segmentation output of SimNIBS (https://simnibs.github.io/simnibs/build/html/index.html), which uses the gray matter surface output from Freesurfer (http://surfer.nmr.mgh.harvard.edu), in 3D Slicer (https://www.slicer.org/) using a closing (fill holes) smoothing method with a 24 mm kernel to fill sharp corners and holes smaller than the kernel size. In cases of interhemispheric subdural electrodes, right and left hemisphere segmentations from Freesurfer are generated and processed in FSL (FMRIB Software Library) through hole filling and mean filtering using a Gaussian kernel of 0.1 mm. For interhemispheric subdural/surface electrodes, we also restricted the sampling of gray matter to the side of the brain the contact recorded from so that the spherical volume would not include gray matter coverage on the opposite hemisphere (Figure 1E).

For each electrode centroid, spheres ranging in radius of 1 mm to 15 mm at 0.5 mm intervals were created to represent the recording volume around each contact (Figure 1F). This recording distance, which we refer to as the radius of influence (RoI), symbolizes the distance away from the contact centroid at which a signal from gray matter can be “recorded” by the electrode. Regarding the coverage of white matter, from which recording is typically attenuated, we interpret the RoI as the volume at which a region may be electrically stimulated, similar in concept to the volume of tissue activated that has been employed in the deep brain stimulation field for many years [18–20]. The probability map generated in LeGUI defines the likelihood that each voxel is gray matter, white matter, cerebral spinal fluid, skin, or bone. To quantify gray and white matter coverage, the volume per voxel is multiplied by the probability each voxel is gray or white matter via fslmaths in the FSL software package (Figure 1F). In some instances, the RoI includes cerebellar gray matter or subcortical regions. In such cases, we did not include these regions in the total gray or white matter quantification to avoid an overestimation of coverage since the use of subcortical and cerebellar recordings in the context of epilepsy are specialized and rare.

To address the concern that spherical RoI may not be an appropriate approximation of actual recording volume, for 3 patients (1 subdural strip patient, 1 subdural strip and subdural grid patient, 1 depth only patient), we created a computational finite element model (FEM) using patient-specific imaging. The FEM voltage solution takes into account the lead geometry (subdural electrode: circle with 2.3 mm diameter; depth electrode: cylinder with height 2.29 mm and diameter of 0.86 mm) and the unique brain geometry of the gyri and sulci on the cortical surface. A tetrahedral head mesh for each patient was created in SCIRun 5 (Scientific Computing and Imaging Institute (SCI), http://www.scirun.org), including cerebral spinal fluid (conductivity=1.79 S/m), gray matter (conductivity=0.33 S/m), and white matter (conductivity=0.142 S/m) as defined in the mesh output of SIMNIBS. The finite element solution, which determines the spread of a voltage signal from an intracranial electrode, was solved at 1 mA stimulation amplitude for all intracranial electrodes using patient-specific tetrahedral head meshes. The current return in the finite element model was defined as the top 5% of the outer cerebral spinal fluid boundary to mimic the use of a series of screws at the top of the head during intracranial cases as ground. FEM-based recording volumes were determined by generating an isosurface at the average voltage along 2.5 mm, 5 mm, 10 mm, and 15 mm spheres around the contact center. This method created a similarly sized volume to the spherical recording volume; however, it accounted for deviations of voltage spread due to the contact and brain geometries. FEM-based recording volumes were compared to spherical volumes using the Dice coefficient, which quantifies overlap and penalizes differences between the two volumes. Finally, gray matter coverage was calculated using FEM-based recording volumes to compare against gray matter coverage using spherical recording volumes at 2.5, 5, 10, and 15 mm.

## Results

This analysis contained 65 patients who underwent intracranial monitoring for intractable epilepsy at the University of Utah hospital from 2010-2020. The mean age (and standard deviation) of patients was 33.4 (10.1) years and the average duration from first seizure until the first monitoring was 19.6 years. The depth and strip electrode cohorts were primarily bilateral whereas the strip and strip + grid cases were typically unilateral (Table 1).

The true volume of recorded tissue for a single electrode contact is highly dependent on signal characteristics, such as neuronal population synchrony and EEG frequency [21,22]. Therefore, a single recording radius cannot apply to the diverse range of potential recording scenarios. Because the volume of tissue recorded depends on both electrode geometry and intrinsic signal properties, we performed an inclusive analysis across a range of biophysically plausible RoIs.

We quantified the amount of gray and white matter coverage normalized by the total number of contacts per patient (Figure 2). Non-normalized gray and white matter coverage can be found in Supplementary Fiigure 1. At RoIs smaller than 4 mm, depth electrodes covered significantly more gray matter than surface electrodes. This increased coverage at small distances using depth electrodes reverses from approximately 4 mm to 12 mm where subdural grid electrodes demonstrated increased gray matter coverage than depth electrodes. With surface electrode modalities, the strip + grid electrode combination covered more gray matter than the strip only configuration at RoIs less than 8 mm; however, strip electrodes cover more gray matter than all other modalities with RoIs greater than 8 mm. Interestingly, at greater than 12 mm RoIs, depth electrodes provided more gray matter coverage than strip + grid and strip + grid + depth hybrid cases. For all values of the RoIs we tested, white matter coverage was much greater in cases that exclusively used depth electrodes, which will be important when considering applications requiring stimulation of white matter tracts to modulate networks [23,24].

**Figure 2.**
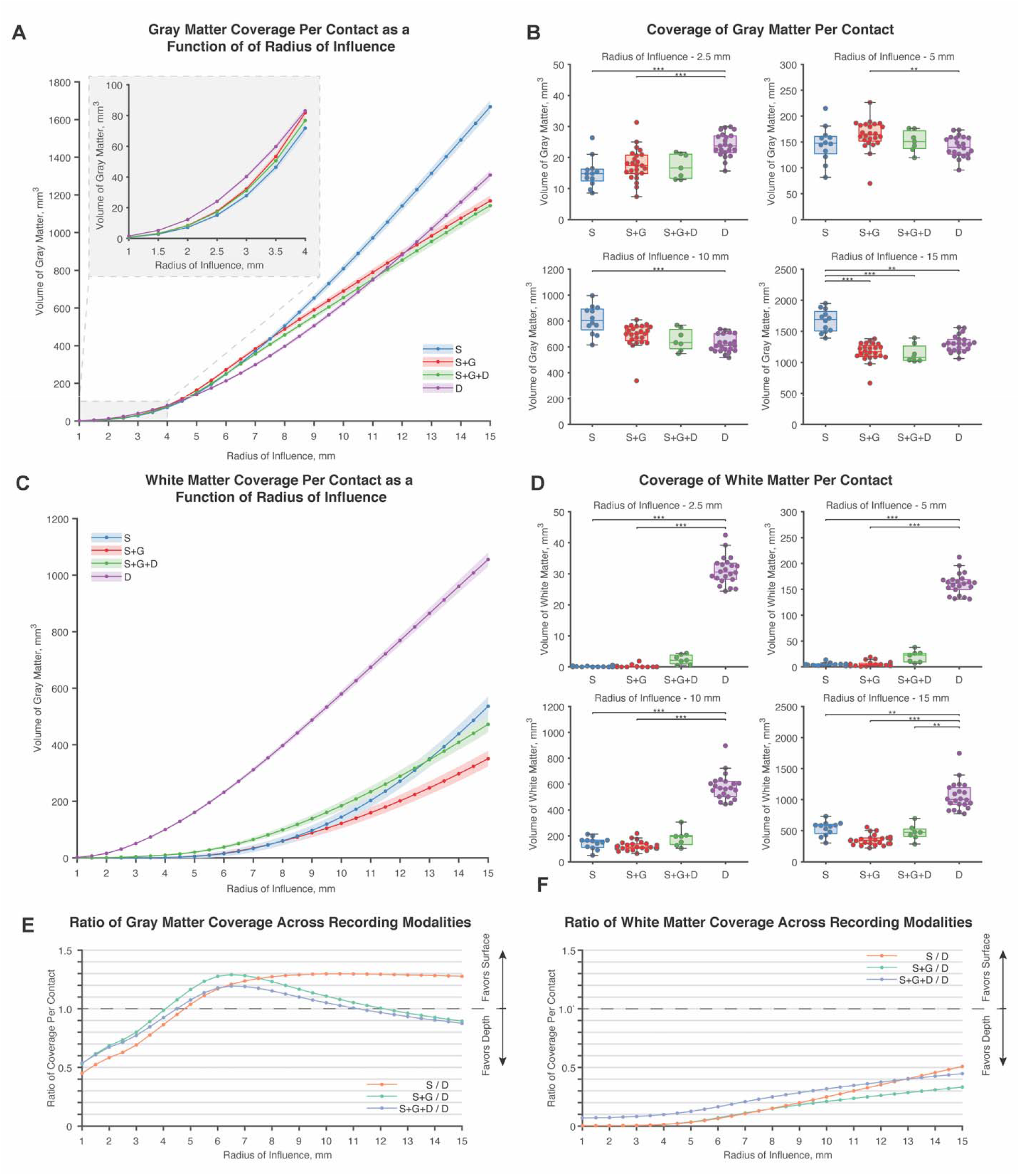
Gray and white matter coverage per contact across RoIs. **A**. There was a dynamic relationship between gray matter coverage and the size of the RoI used across intracranial modalities. **B**. Acros notable RoIs at 2.5 mm, 5 mm,10 mm, and 15 mm, depth electrodes covered more gray matter at small RoIs than strip electrode or strip and grid electrode combinations (S & D: p = 3.2 × 10^−5^ ; S+G & D: p =5.34 × 10^−5^); however, the relationship reversed for strip + grid and depth electrodes at a 5 mm RoI (S+G & D: p = 0.00326) and reversed for strip and depth electrodes at a 10 mm RoI (S & D: p = 1.86 × 10^−5^). At 15 mm, strip electrodes exceeded other intracranial electrode approaches in gray matter coverage on a per contact basis (S & S+G: p = 5.17 × 10^−7^ ; S & S+G+D: p = 3.60 × 10^−5^; S & D: p = 0.00477);. **C**. White matter coverage varied across recording modalities, but was dominated by depth electrodes. Though not significant, cases that used strip, grid, and depth electrodes in combination appeared to increase the white matter sampling over cases that used subdural electrodes alone at RoIs less than 12 mm. **D**. Depth electrodes sample more white matter than configurations that used subdural electrodes. (2.5 mm - S & D: p = 1.61 × 10^−5^ ; S+G & D: p = 3.84 × 10^−9^; 5 mm - S & D: p = 1.94 × 10^−6^ ; S+G & D: p = 4.45 × 10^−9^; 10 mm - S & D: p = 9.58 × 10^−5^ ; S+G & D: p = 4.02 × 10^−9^; 15 mm - S & D: p = 0.00768; S+G & D: p = 3.78 × 10^−9^; S+G+H & D: p = 0.00446) **E**. Relative to depth electrodes, subdural electrodes covered less gray matter prior to a 4 mm RoI. At larger radii, subdural electrodes covered more gray matter until a RoI of 12 mm when only the depth electrodes surpassed configurations strip and grid configurations and strip, grid, and depth configurations. **F**. Across all RoIs, depth electrodes covered more white matter. White matter coverage for very small RoIs was close to zero, while at very large RoIs, depth electrodes covered approximately twice the volume of white matter as subdural electrodes.

We quantified region-specific gray matter coverage in the frontal lobe, temporal lobe, hippocampus, amygdala, insula, and cingulate cortex using the macroscopic labels from the AAL atlas. Patients were included in the region-specific analysis if there was any gray matter coverage in that region at a 5 mm RoI. Patients who did not meet this standard were excluded as the region in question was likely not included in the localization hypothesis.

Regardless of localization hypothesis, all patients had gray matter coverage in the frontal and temporal lobes (Figure 3A). Patients with a combination of subdural strip and grid electrodes surpassed gray matter coverage in the temporal lobe compared to cases that used subdural strip electrodes alone (p = 2.27 × 10^−4^).

**Figure 3.**
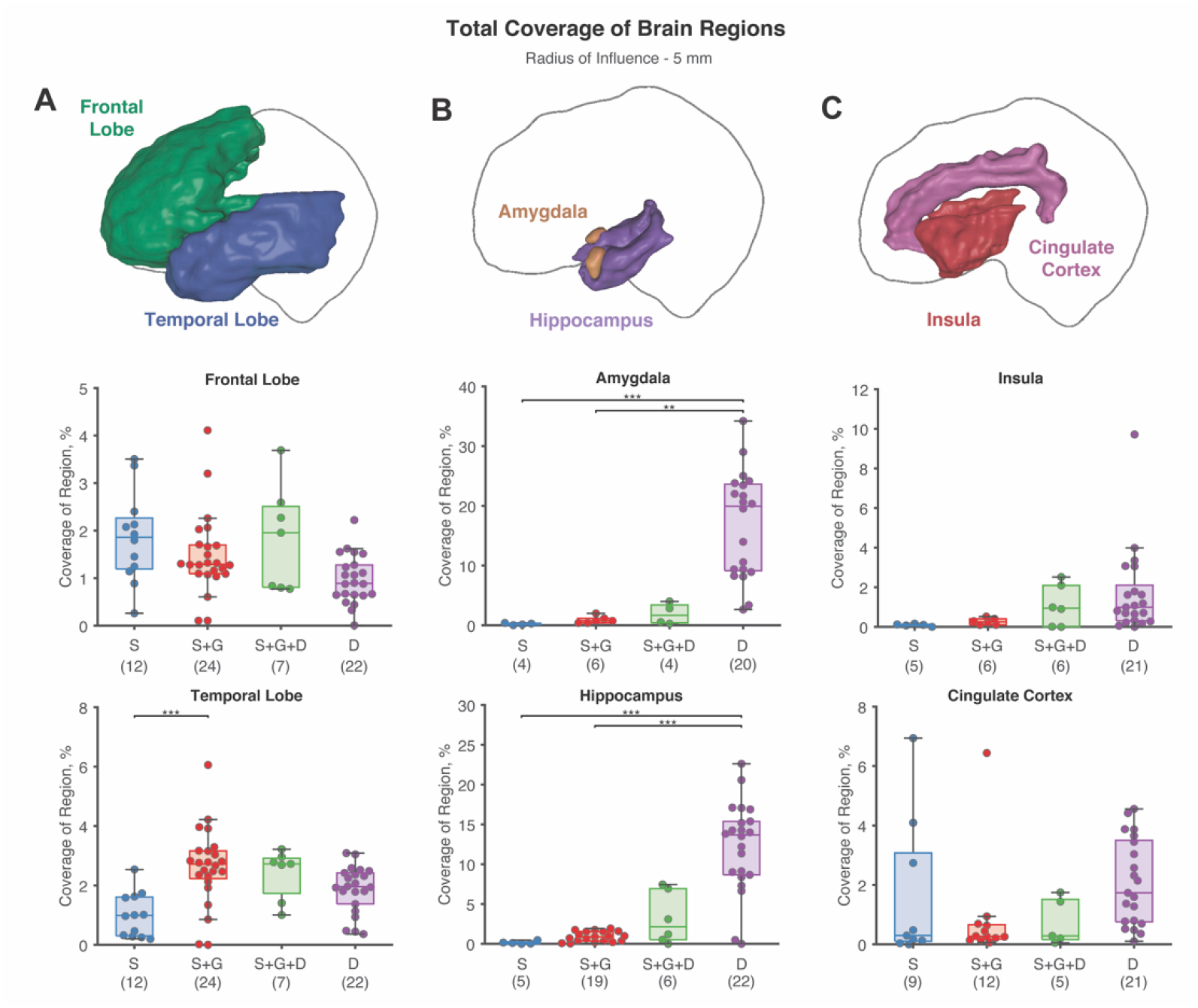
Region-specific coverage of gray matter was calculated across all patients. (n) on the x-axis label indicates the number of patients included. **A**. All patients had some coverage of the frontal and temporal lobes regardless of the modality used. **B**. Notably, hippocampal and amygdala coverage of depth electrodes greatly exceeded coverage using subdural electrode configurations. **C**. Patients with depth contacts only had significantly more insula coverage than patients with subdural strip contacts. Nearly all depth electrode only patients had coverage in the insula and cingulate cortex.

Mesial temporal lobe structures such as the hippocampus (S & D: p = 1.17 × 10^−4^; S+G & D: p = 4.574 × 10^−5^) and the amygdala (S & D: p =6.33 × 10^−4^; S+G & D: p = 0.00453) had significantly more coverage in depth electrode cases than cases with subdural strips only or a combination of subdural strips and grids (Figure 3B). While hybrid cases that used both subdural electrodes and depth electrodes appeared to increase coverage of the hippocampus and amygdala, there were not enough patients to establish significance. Finally, we quantified gray matter coverage in additional limbic structures including the insula and cingulate cortex (Figure 3C). No modality covered significantly more gray matter in the insula or cingulate cortex than others, however, it should be noted that nearly all (21/22) depth patients had some coverage in both of these regions, independent of localization hypothesis. In contrast, only 5/12 patients with strip electrodes and 6/24 patients with strip and grid electrodes had coverage in the insula, while 9/12 patients with strip electrodes and 12/24 patients with strip and grid electrodes had coverage in the cingulate cortex.

Three patient-specific computational FEMs were generated to explore an alternative method to the spherical RoI in estimating the recording volume. FEM solutions were computed for 1 mA for all electrodes (Figure 4A). FEM-based recording volumes that followed the equipotential lines of the voltage solution were compared to the spherical volumes using the Dice coefficient (Figure 4B), in which a value of zero indicates no similarity while a one indicates perfect overlap. Median Dice coefficients across all RoIs were 0.845, 0.867, and 0.919 for the strip, strip and grid, and depth patients, respectively, and these values indicate that the spherical volumes were reasonably similar to the FEM-based volumes. After a conducting a chi-squared test with a bonferroni multiple comparison correction, FEM-based volumes were not found to be significantly different in shape to the spherical volume in all cases. When calculating the differences in gray matter coverage between spherical volumes and FEM-based volumes, there was no statistical significance in any patient at any of the RoI tested (Figure 4C) which further supports the use of a spherical volume to approximate a contact’s RoI.

**Figure 4.**
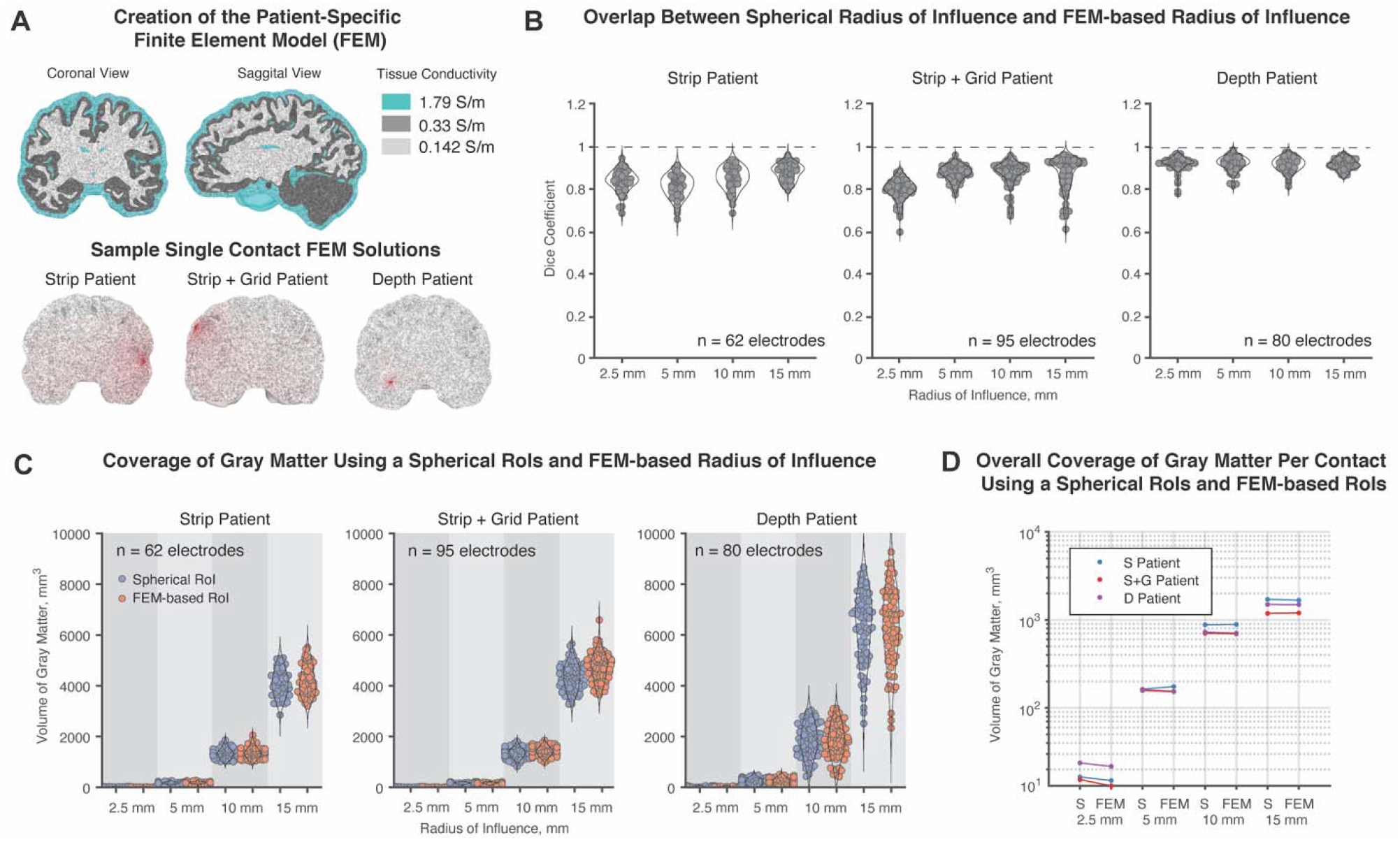
Use of patient-specific brain modeling to create RoIs. **A**. Using a head mesh generated from a FreeSurfer/SIMNIBS pipeline. The finite element method was used to solve for voltage solutions for all electrodes for 3 patients (1 strip patient, 1 strip and grid patient, 1 depth patient). FEM-based RoI volumes were determined by equipotential lines according to the average voltage at 2.5 mm, 5 mm, 10 mm, and 15 mm away. **B**. Using the Dice coefficient to measure overlap, FEM-based volumes were not statistically significantly different in shape to spherical volumes. **C**. Slight differences in individual FEM-based volumes did not translate to significant differences in the calculated coverage of gray matter. **D**. Overall, across patients and all RoIs, there was no consistent pattern in how total gray matter coverage changed with the use of a FEM-based RoI, thus, justifying the use of spherical RoIs in our model.

## Discussion

Despite decades of clinical use, it is unclear whether subdural grid and strip electrodes record from more cortical gray matter than penetrating depth electrodes in normal clinical settings. This analysis may contribute to choosing a modality for seizure localization during intracranial monitoring, and the goal of this study was to quantify gray and white matter coverage through patient-specific imaging and probabilistic segmentations.

In this study, we used the radius of influence, RoI, to define the recording distance of an electrode. As in any computational model, it is important to determine the sensitivity of an outcome metric to specific parameters in the study. For this reason, we varied the RoI from 1 mm to 15 mm in 0.5 mm steps and calculated the total volume of gray and white matter covered at each RoI. In doing so, we found that the choice of RoI greatly impacts the amount of gray matter sampled across intracranial electrode modalities. Most notably, at a 2.5 mm RoI, gray matter coverage using depth electrodes exceeded the gray matter coverage using either strip electrodes or a strip and grid electrode combination. This relationship held in gray matter coverage per contact (Figure 2B) as well as total gray matter coverage (Supplementary Fiigure 1A). Strip electrodes exceeded depth electrodes in gray matter coverage at both 5 mm and 10 mm RoIs; the combined use of strip and grid electrodes exceed depth coverage at a RoI of 10 mm only. This reversal in gray matter coverage is likely due to the typical intercontact spacing of subdural and depth electrodes. With the exception of a few patients, most depth leads in this study had a 5 mm intercontact spacing while all subdural electrodes in this study had a 10 mm spacing. When the RoIs exceeded 2.5 mm for depth electrodes, they overlapped with recording volumes from neighboring contacts, thereby creating redundancy in gray matter coverage. This intercontact spacing explains why at greater RoIs, specifically greater than a 4 mm RoI, subdural electrodes sampled more gray matter. Beyond an 8 mm RoI, strip electrodes sampled more gray matter, presumably because strip subdural electrodes have the least amount of redundancy across all the modalities we tested.

Tantawi et al. recently reported gray matter coverage using a fixed radius of 2.5 mm. They included 20 patients, with 10 patients in the depth cohort and 10 in the surface electrode cohort Patients in this study were not separated into separate cohorts based on the use of subdural grid electrodes versus strip-only configurations. The analysis found that, using a single spherical recording volume with a radius of 2.5 mm, the average per contact coverage was not significantly different across the depth and surface modalities, though depth electrodes sampled more total gray matter volume than surface electrodes. Using a comparable 2.5 mm RoI in our study, we found that depth electrodes sampled significantly more gray matter than strip and strip + grid combinations in per contact (Figure 2B) and total coverage calculations (Supplementary Fiigure 1A). The discrepancy across our results and previously reported values is likely due to our use of probabilistic calculations of gray and white matter which offer more precise parcellation of gray and white matter regions within each voxel. Additionally, the majority of SEEG leads in our study used 5 mm intercontact spacing, while Tantawi et al. (2021) used SEEG leads with 3.5 mm intercontact spacing. We found it critical to separate strip and strip + grid modalities in our analysis because, at RoIs greater than 8 mm, strip only configurations sampled more gray matter than strip + grid configurations (Figure 2A). It should also be noted that Tantawi et al. (2021) had larger electrode counts than in our analysis, which may explain why depth electrodes sampled significantly more gray matter in the total gray matter calculation but not in the per contact volume calculation. The mean (and standard deviation) of electrode contacts for our study was 92.8 (18.5) depth, 74.6 (16.2) strip, and 86.4 (22.8) strip + grid electrodes while Tantawi et al. (2021) reported 192.1 (54.2) depth, 128 (49.6) strip, and 110.4 (36.1) strip + grid electrodes. Finally, our study also included a sensitivity analysis to test how the assumed RoI, ranging from 1 to 15 mm, could alter the gray matter coverage results and found the choice in RoI can greatly influence which modality reportedly records more gray matter.

In the context of white matter, the RoI in this study can be interpreted similarly to the volume of tissue activated, which quantifies the extent of activation in tissue in response to electrical stimulation of a contact. While we do not use an estimate of activation as described by classic methods of volumes of tissue activated, volumes of tissue activated are roughly spherical and have an approximate radius of 2-5 mm from the contact centroid for standard deep brain stimulation waveforms (pulse width, 60-90 µs) and increase in size with increasing amplitude and pulse width. [18,19,26,27]. While the finding that depth electrodes sample more white matter may not be surprising, it may be important to quantify white matter coverage given that intracranial stimulation procedures, such as ictal mapping and sensorimotor mapping, need to map white matter tracts to fully interrogate involved circuits. A recent study on ictal mapping using SEEG has shown that a greater percentage of patients with good surgical outcomes had stimulation induced seizures compared to the poor outcome cohort [28]; a finding that motivates the use of depth electrodes to interrogate seizure networks. The utility in quantifying white matter coverage may also be relevant in patients who are candidates for neuromodulation therapy to control their seizures using responsive neurostimulation, as recent evidence indicates that placement of leads and stimulation in the parahippocampal or temporal stem white matter may more effectively target the epileptic network in TLE without sacrificing recording ability [29].

### Region-specific coverage

Coverage of the frontal and temporal lobes was seen in all patients in our study, regardless of the localization hypothesis. However, despite the coverage of these regions in all patients, only a small fraction of the total frontal and temporal lobes were sampled, about 2% (Figure 3A). This is particularly striking because even though subdural grids visibly cover large regions of the cortical surface, the overall regional coverage was comparable to depth electrodes. Unsurprisingly, depth electrodes offered superior coverage of the amygdala and hippocampus that was not achievable in cases that used subdural strips or a combination of subdural strips and grids. This finding motivates the use of depth electrodes for patients with suspected mesial temporal lobe epilepsy, the most common type of focal epilepsy [30]. Nearly all the depth electrode cases had some coverage of the insula or cingulate, while many patients with subdural electrodes were excluded from the analysis because they had no coverage in those regions and, thus, did not meet the criteria for the analysis. Though insular and cingulate cortex epilepsies are rare, nonlesional insular and cingulate epilepsies are difficult to diagnose and may be misdiagnosed as temporal lobe epilepsy or frontal lobe epilepsy without utilization of depth electrodes in those regions [31,32]. Depth electrodes therefore provide a significant advantage in sampling the insula and cingulate, even if those regions may not be a part of the initial localization hypothesis.

### Which radius of influence is the right approximation of recording coverage?

It is unlikely that a single recording distance is valid for all brain areas and states—this distance likely changes based on the number and level of synchrony of cells as well as whether they follow an ordered or irregular arrangement relative to the electrode [33,34]. Recorded local field potentials are largest in the cortex given the compounding of dipoles from active neurons due to the regularized arrangement of neurons in a gyrus and the close proximity of apical dendrites to the electrode surface. This pattern is in contrast to the less regular arrangement of neurons in subcortical structures [22]. However, even within the cortical surface, the folds of a sulcus, in which the highly ordered cell layers bend down and away from surface electrodes, add spatial irregularity that may reduce the size of the signal. Signal strength decays with distance from the source, and on top of that, the cancellation of dipole signals increases with distance as well. [35]. For depth contacts, many contacts are in close proximity to white matter based on findings from Figure 2C-D. Because white matter has very little recordable signal compared to gray matter [36], dipole cancellation of a gray matter signal recorded by contacts located inside or at the boundary of white matter may be reduced, enabling detection at greater distances.

In general, using a spherical RoI is a common way to demarcate regions of coverage around an electrode contact. In analyses used to correlate resting-state activity from intracranial recordings with fMRI, authors have previously used a 6 mm RoI [37] and a 5 mm RoI [38]. According to Dubey and Ray 2019, spatial spread for ECoG electrodes—sized identically to those used in our study—was minimally larger than the size of the electrode itself, at approximately a 3 mm RoI. Finally, scalp electrodes are able to record intracranial activity, despite being more than 1cm from the presumed sources. Ultimately, it’s possible that an appropriate RoI to record a seizure may well be greater than previously reported in literature focused on modeling the recording of normal electrical activity since the level of synchrony is greatly increased across substantial populations of neurons throughout the brain during a seizure.

Finally, the distance at which a signal may be recorded does not appear to be constant across all frequencies. For gamma activity, which is largely thought to represent the firing of local populations of neurons, Szurhaj et al. 2005 reported that gamma activity on one contact did not appear on contacts 1.5 mm away. Correlation of neural activity between electrode pairs can be used as a metric of redundancy across contacts, meaning that the same signal has reached contacts in different locations. A study by Muller et al. 2016 found that such paired correlations could be found across contacts up to 20 mm apart for lower frequencies bands such as theta and alpha. This indicates that some low frequency signals may have sources at great distances away, and supports the reality that even non-invasive scalp electrodes can still record fields from the brain at great distances away, similarly at lower frequency bands [21,42]. In contrast, in the high gamma and gamma frequency bands, correlations between pairs of contacts 5 mm away show markedly reduced correlations. Taking this information into account, it may be valuable to consider signal characteristics when choosing a modality in addition to weighing other factors such as the localization hypothesis or complication risks. Specifically, given that gamma frequency is often recorded closer than 4 mm away from an electrode, depth electrodes appear to be more promising to detect gamma signals, while slow waves, such as alpha and theta, may be more detectable using subdural electrodes given the increased gray matter coverage at 4 to 12 mm distances away from contacts.

### Limitations

The primary limitation in this study includes the use of spherical RoIs to model electrode coverage of gray and white matter regions. In figure 4, we sought to address this limitation by building patient-specific head models for a strip electrode patient, strip and grip electrode patient, and a depth electrode patient to generate FEM-based recording volumes. The advantage in taking a patient-specific approach is that the estimated recorded volume can account for the contact geometry and how different conductivities of cerebral spinal fluid, gray matter, and white matter can alter the spread of the voltage signal through the brain. Despite the difference in shape of the recording volumes, the amount of gray and white matter covered by each contact was not significantly different across different RoIs for all patients in the FEM-based approach (Fig 4C). Additionally, in order to have a cohort of 65 patients, we did not exclude patients based on their clinical localization hypothesis. It may be valuable in future work to compare gray and white matter coverage across modalities in patients with similar localization hypotheses and determine if there are region-specific trends in gray and white matter coverage. Another limitation is the restriction of our cohort to a single center. Though this cohort is drawn from two neurosurgeons with different surgical approaches, it is possible that other centers may place electrodes in such a way as to change the precise points at which depth electrodes outperform surface electrodes. Lastly, the surface electrodes used in this study come from a single manufacturer and have a consistent inter-contact distance. Other manufacturers or other electrode spacings may affect these results, as well.

## Statistical Methods

Statistics were calculated in MATLAB. To avoid the assumption of normality in our dataset, significance across groups was calculated using the non-parametric Kruskal-Wallis test. In all cases of multiple comparisons, a Bonferroni correction was applied and p-values less than 0.05 were considered statistically significant. For Dice coefficient calculations, we used the Pearson’s chi-squared test at the p<0.05 with a Bonferroni correction to determine whether FEM-based and spherical radii of influence significantly differ in volume to the spherical radii of influence.

## Data Availability

Data used in this study are available upon reasonable request.

## Contributorship

DNA completed the data analysis and wrote the first draft of the manuscript. CMC built patient specific computational head models. EHS provided guidance on the data analysis and statistical methods. AMA contributed retrospective patient data. TDS created the LeGUI software and provided guidance on atlas registration and segmentation. JDR provided project guidance. All authors contributed to the editing of the final manuscript.

## Ethics approval

Retrospective analysis of the data is approved by the University of Utah Institutional Review Board under exemption #115230.

