## Supplementary for "Probabilistic comparison of gray and white matter coverage between depth and surface intracranial electrodes in epilepsy: a patient-specific modeling and empirical study"

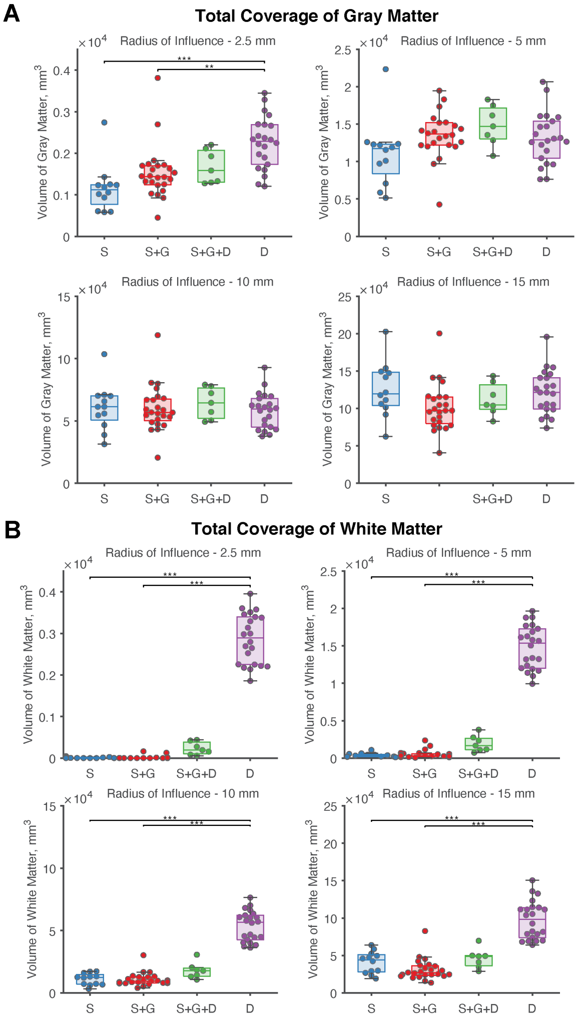


Supplementary Figure 1. Analysis of total gray and white matter coverage without normalization based on number of contacts implanted. **A.** At 2.5 mm RoI, depth electrodes have significantly greater coverage than cases using subdural strip electrodes or a combination of subdural strip or grid electrode configurations. This significant relationship is not seen at 5 mm, 10 mm, and 15 mm RoIs. **B.** For all RoIs shown, depth electrodes cover a greater amount of white matter modalities that only use subdural electrodes.

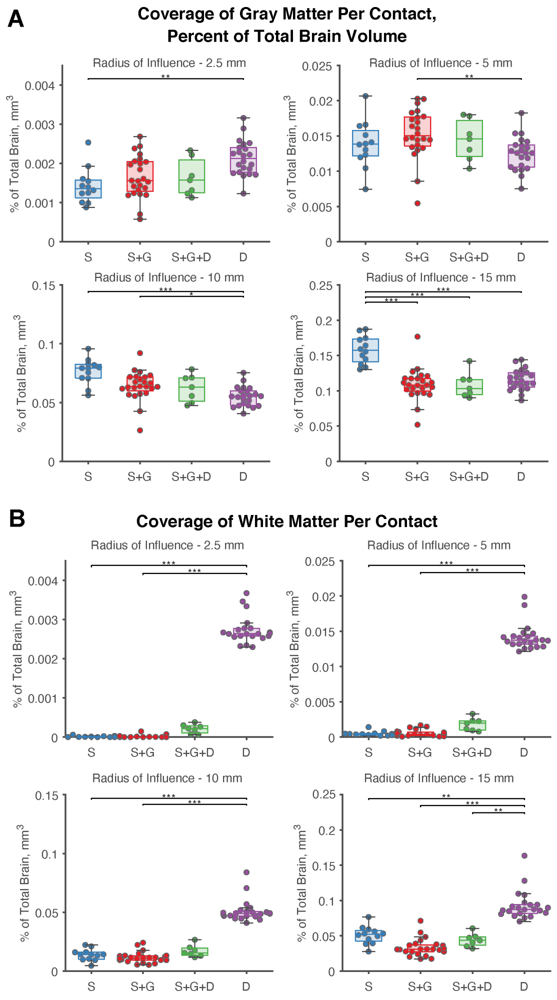


Supplementary Figure 2. Per contact coverage as a percent of total brain volume across all recording configurations. **A.** Gray matter coverage is greatest for depth electrodes at 2.5 mm compared to strip electrodes. Strip electrodes cover the most gray matter compared to all other modalities at 15 mm. For a RoI of 5 mm, electrodes each cover approximately 0.01-0.02% of total brain coverage. As RoI increases, the percentage of brain coverage increases with diminishing returns seen likely due to redundancy. **B.** Similarly, for all RoIs shown, depth electrodes cover a greater amount of white matter per contact than cases using subdural electrodes.
